# Canadian Healthcare Providers’ Attitudes Towards Automated Insulin Delivery Systems

**DOI:** 10.1101/2022.06.02.22275169

**Authors:** Amy E Morrison, Kate Farnsworth, Holly O Witteman, Anna Lam, Peter A Senior

## Abstract

**Introduction:** We aimed to assess the current experience and attitudes towards Commercial and Do-it-yourself (DIY) automated insulin delivery (AID) systems among healthcare providers (HCP) across Canada.

**Methods:** A cross-sectional study was performed through electronic distribution of an anonymous survey to HCP licensed to practice in Canada looking after people with type 1 diabetes (T1D).

**Results:** Responses included 204 HCP across the multi-disciplinary team; dieticians (32.8%), nurses (31.9%), and endocrinologists (28.4%), looking after adults (51%) and children (23%) mainly in urban areas (85.7%). Respondents reported a median 100-500 patients with T1D per practice, with a median 6-24 current users/practice of Commercial compared to a median 1-5 current users/practice of DIY AID. The majority of HCP (72.7%) were comfortable supporting Commercial AID, whereas only 21.6% reported comfort supporting DIY AID use. A significant, although moderate correlation between HCP experience and comfort was seen; Commercial r=0.57(p<0.0001) and DIY r=0.45(p<0.0001). Respondents reported more barriers to DIY, relative to Commercial AID(p=0.001); unfamiliarity/lack of exposure and medico-legal risks were highlighted with DIY systems. Respondents suggested AID system education (both Commercial and DIY), for HCP and users, to improve HCP confidence.

**Conclusions:** Despite documented beneficial outcomes, AID systems are not widely used in the management of T1D in Canada. The need for both user and HCP education to improve familiarity with the systems, in addition to clarity in medico-legal guidance, have been identified as gaps, which if addressed, might enable the benefits of AID to be more widely available to people with T1D in Canada.

## Introduction

Automated Insulin Delivery (AID) or closed-loop systems combine an insulin pump with continuous subcutaneous insulin infusion (CSII), and a Continuous Glucose Monitor (CGM), controlled by a computerized predictive algorithm. This enables automated adjustment in insulin delivery rate based on CGM data. These systems are termed ‘hybrid closed-loop’ due to the integration of this automated, algorithm mediated insulin delivery, with additional user input, such as mealtime insulin boluses [1]. Commercial AID systems are associated with improved glycemic outcomes and are the most technologically advanced, regulatory approved method of insulin delivery [2].

In Canada, Commercial AID was first introduced in 2016 and three systems are currently available; Medtronic 670G and 770G (combining a Medtronic pump and Guardian 3 sensor) and Tandem Control-IQ (combining Tandem t:slim X2 pump and Dexcom G6 sensor) [3]. Despite some clear beneficial outcomes, notably in glycemia, quality of life and safety, these systems are felt to be suboptimal by many people with type 1 diabetes. The systems are expensive, development and incorporation of new features is a slow process, glucose targets lack flexibility, and therefore these systems do not meet the lifestyle needs of many users. In contrast, novel, unregulated and unapproved, user-designed or do-it-yourself (DIY) AID systems are increasingly being used. These systems which were developed prior to the introduction of any Commercial AID systems, are categorized according to the algorithm and technology which they incorporate into; OpenAPS, AndroidAPS and Loop systems. Users build their own DIY AID system with the help of online instructions and support from other users via social-media platforms [4].

The user-built and unregulated nature of DIY AID systems makes them a challenging prospect for healthcare providers (HCP) caring for people with type 1 diabetes that are currently using or contemplating commencing use of one of these systems.

Approved technologies (CSII and CGM), prescribed by HCP, are effectively being ‘hacked’ and implemented in an unregulated and unapproved way by the user [5]. There has been no official guidance for HCP in Canada as to how they should address patients who are using, or planning to use a DIY AID system.

HCP opinions and current practices towards DIY AID use have previously been collected in studies in both the United Kingdom (UK) [6] and United states of America (USA) [7,8]. The UK study, used an online questionnaire to survey the opinions of 317 HCP (46% consultants and 8% registrars/trainees in Diabetes and Endocrinology, 38% diabetes specialist nurses or dieticians and 8% other HCP). One cross-sectional study from the USA, used a paper-based survey to collect opinions from 47 HCP (90.7% female), in addition to an online survey, evaluating usefulness and acceptability of an AID education and a comparison factsheet, with 137 responses (93% female), 91% of these respondents were diabetes nurses and nutritionists [7]. A second study from the USA reported an American Association of Diabetes Educators HCP survey with 152 respondents, of these 27% reported that they felt DIY AID systems were safe [8], with just 2% of the UK HCP respondents perceiving DIY AID as dangerous [6].

We performed a cross-sectional to study to assess current HCP knowledge, experience and attitudes towards AID across Canada. We aimed to highlight prevalent areas of knowledge gaps and consistent patterns in HCP concerns in order to direct future targeted HCP education and consensus guidelines, to ultimately ensure that users of AID systems receive consistent and appropriate patient care.

## Methods

A 31-item anonymized online survey was designed, the development process involved both assessment of face validity from HCP, as well as perspectives from patient-researchers active in the DIY community. The final version of this survey (appendices figure 1.) was estimated to take approximately twenty minutes to complete, and was distributed using the REDCap (Research Electronic Data Capture) system. REDCap is a secure, web-based software platform, designed to support data capture for research studies [9,10]. Participants were HCP licensed to practice in Canada, looking after children and/or adults with type 1 diabetes. The survey comprised sections relating to; HCP and practice characteristics, HCP current experience and attitudes towards AID, perceived barriers to AID use, comfort levels with AID use in specific patient scenarios and potential enablers of AID. Participants were asked to answer all questions, with the option of not applicable (n/a), unsure, or prefer not to say always available. An additional six questions relating to specifics of DIY AID system settings and applications used in conjunction with DIY AID systems, were asked to respondents that deemed themselves to be ‘actively involved’ in the care of people using DIY AID systems.

The study was approved by the University of Alberta Research Ethics Board, study ID Pro00108472. A snowball sampling strategy was employed. An electronic link to the survey was sent to prospective participants through their place of work, diabetes specialist networks in Canada and additionally distributed via social media platforms. Participant responses were anonymous, with no directly identifiable information in responses, the only potentially identifiable data related to province and setting of practice (whether rural or urban, and in a community or academic centre).

Descriptive statistical analysis, Spearman correlation coefficient, paired Wilcoxon signed rank test (with comparison of HCP responses relating to the two system types) and Kruskal-Wallis test with Dunn’s multiple comparison test (for subdivision of responses according to HCP type), were performed using GraphPad Prism version 9.2.0 for macOS; GraphPad software, San Diego, California, www.graphpad.com. The level of statistical significance was defined as a p value <0.05. NVivo 12 QSR; Melbourne, Australia, www.qsrinternational.com, was utilized to identify the dominant issues in the HCP qualitative responses, through word count and word cloud analysis [11].

## Results

### HCP and Practice Characteristics

In total, n=204 responses were collected with the online survey open for 35 days; June 25^th^ until July 30^th^ 2021. HCP practice characteristics are shown in table 1. The most prominent locations for respondents were Ontario 75 (36.8%), Alberta 65 (31.9%), Quebec 18 (8.8%) and British Columbia 17 (8.3%) in Community Urban 84 (41.2%), Academic Centres 58 (28.4%) and Urban Hospitals 33 (16.1%). The majority of respondents 104 (51%) care for adults with type 1 diabetes, with 47 (23%) children and 53 (26%) both, with 112 (54.9%) designated Certified Diabetes Educators (CDE).

**Table 1.**
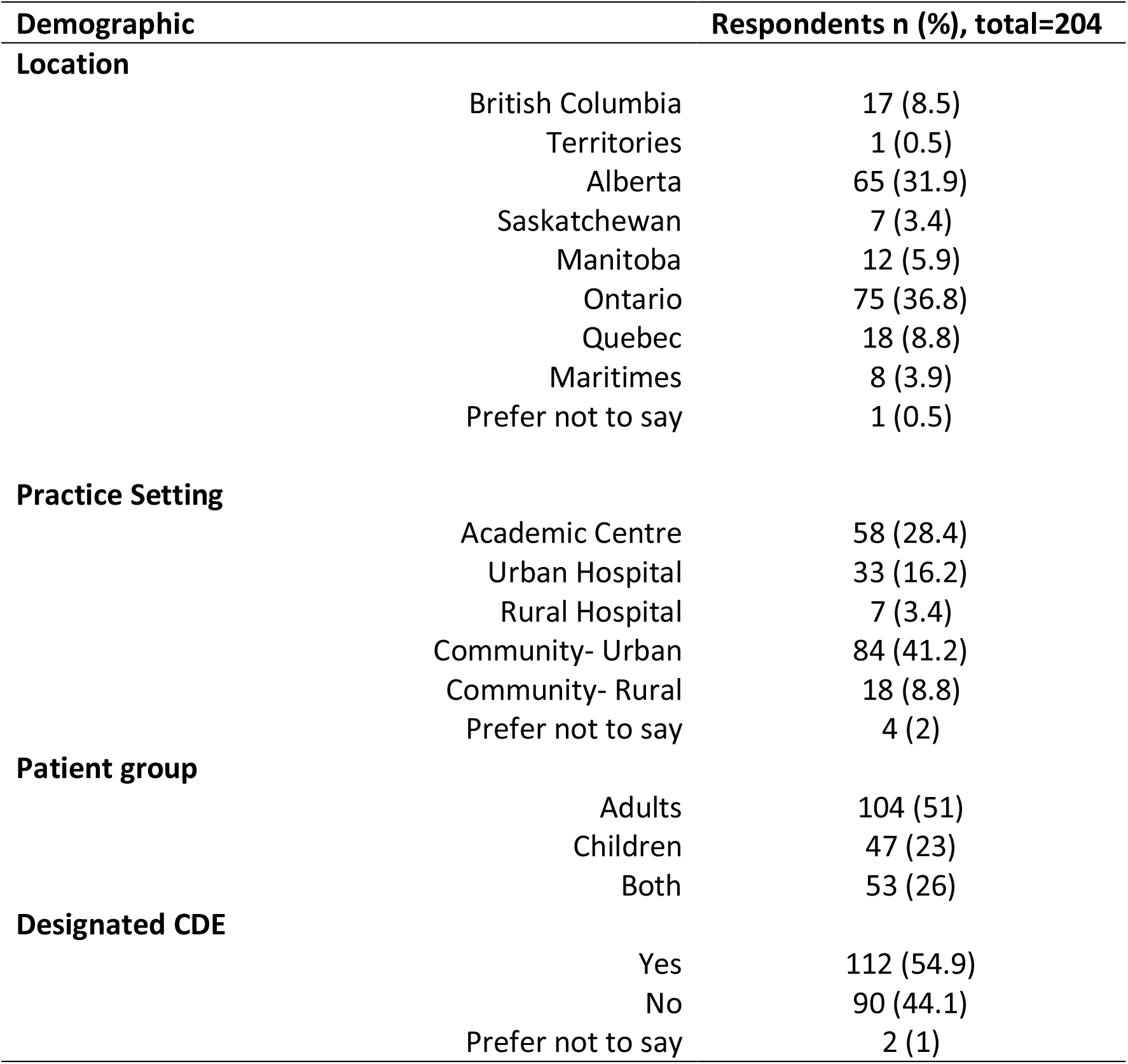
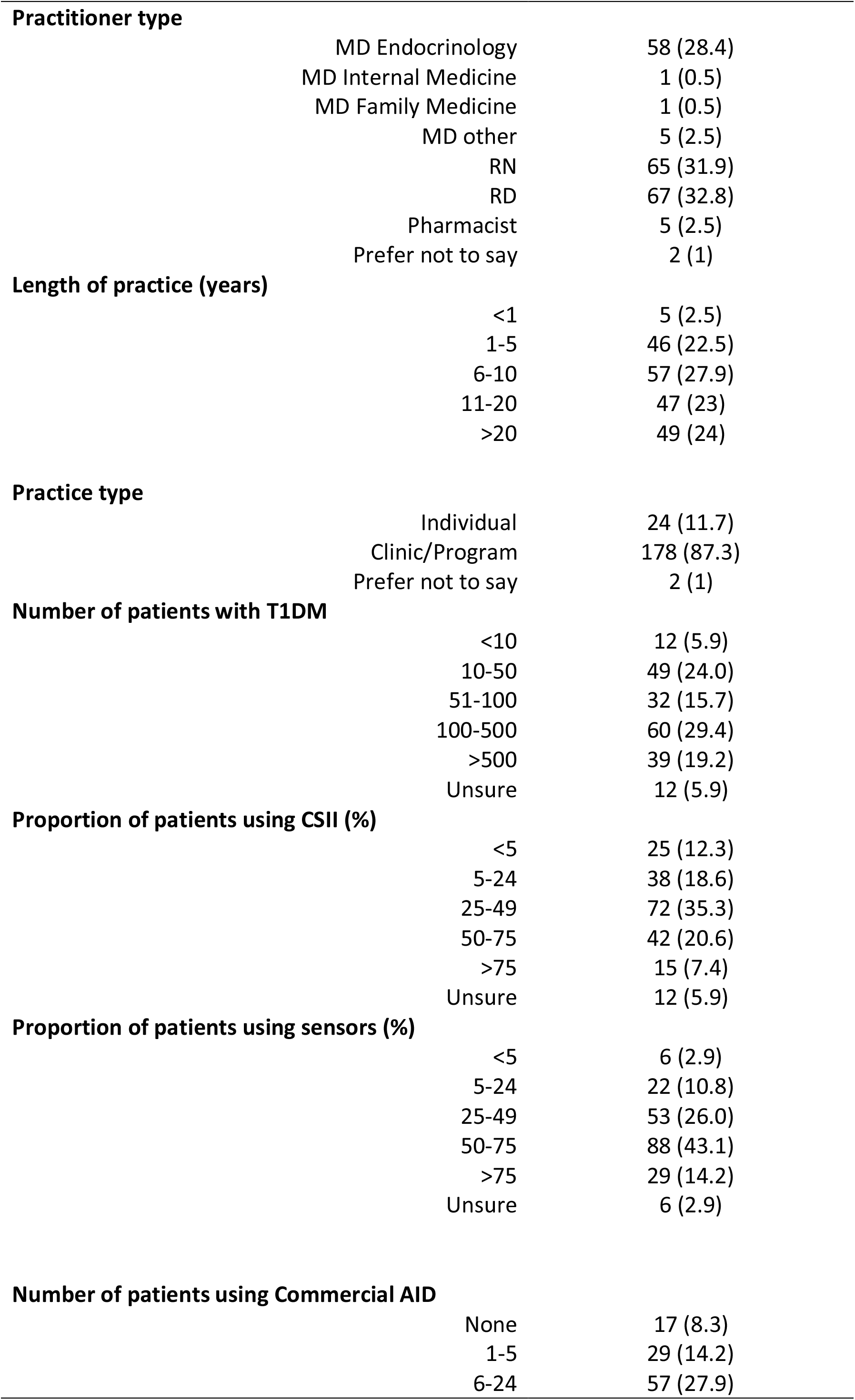
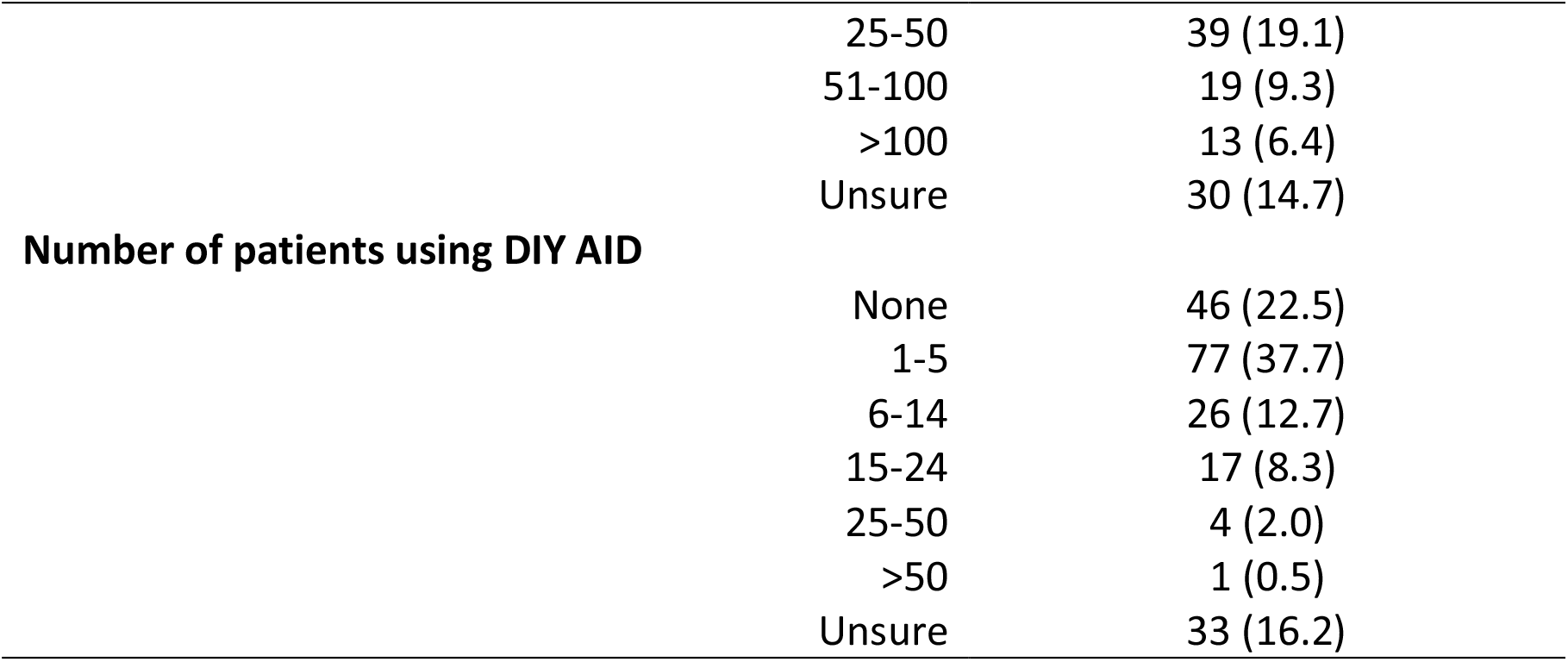
Healthcare Provider Characteristics and Technology Experience of Survey Respondents.

| **Demographic** | **Respondents n (%), total=204** |
| --- | --- |
| **Location** |  |
| British Columbia | 17 (8.5) |
| Territories | 1 (0.5) |
| Alberta | 65 (31.9) |
| Saskatchewan | 7 (3.4) |
| Manitoba | 12 (5.9) |
| Ontario | 75 (36.8) |
| Quebec | 18 (8.8) |
| Maritimes | 8 (3.9) |
| Prefer not to say | 1 (0.5) |
| **Practice Setting** |  |
| Academic Centre | 58 (28.4) |
| Urban Hospital | 33 (16.2) |
| Rural Hospital | 7 (3.4) |
| Community- Urban | 84 (41.2) |
| Community- Rural | 18 (8.8) |
| Prefer not to say | 4 (2) |
| **Patient group** |  |
| Adults | 104 (51) |
| Children | 47 (23) |
| Both | 53 (26) |
| **Designated CDE** |  |
| Yes | 112 (54.9) |
| No | 90 (44.1) |
| Prefer not to say | 2 (1) |

|  |  |  |
| --- | --- | --- |
| **Practitioner type** |  |  |
|  | MD Endocrinology | 58 (28.4) |
|  | MD Internal Medicine | 1 (0.5) |
|  | MD Family Medicine | 1 (0.5) |
|  | MD other | 5 (2.5) |
|  | RN | 65 (31.9) |
|  | RD | 67 (32.8) |
|  | Pharmacist | 5 (2.5) |
|  | Prefer not to say | 2 (1) |
| **Length of practice (years)** |  |  |
|  | <1 | 5 (2.5) |
|  | 1-5 | 46 (22.5) |
|  | 6-10 | 57 (27.9) |
|  | 11-20 | 47 (23) |
|  | >20 | 49 (24) |
| **Practice type** |  |  |
|  | Individual | 24 (11.7) |
|  | Clinic/Program | 178 (87.3) |
|  | Prefer not to say | 2 (1) |
| **Number of patients with T1DM** |  |  |
|  | <10 | 12 (5.9) |
|  | 10-50 | 49 (24.0) |
|  | 51-100 | 32 (15.7) |
|  | 100-500 | 60 (29.4) |
|  | >500 | 39 (19.2) |
|  | Unsure | 12 (5.9) |
| **Proportion of patients using CSII (%)** |  |  |
|  | <5 | 25 (12.3) |
|  | 5-24 | 38 (18.6) |
|  | 25-49 | 72 (35.3) |
|  | 50-75 | 42 (20.6) |
|  | >75 | 15 (7.4) |
|  | Unsure | 12 (5.9) |
| **Proportion of patients using sensors (%)** |  |  |
|  | <5 | 6 (2.9) |
|  | 5-24 | 22 (10.8) |
|  | 25-49 | 53 (26.0) |
|  | 50-75 | 88 (43.1) |
|  | >75 | 29 (14.2) |
|  | Unsure | 6 (2.9) |
| **Number of patients using Commercial AID** |  |  |
|  | None | 17 (8.3) |
|  | 1-5 | 29 (14.2) |
|  | 6-24 | 57 (27.9) |
|  | 25-50 | 39 (19.1) |
|  | 51-100 | 19 (9.3) |
|  | >100 | 13 (6.4) |
|  | Unsure | 30 (14.7) |
| **Number of patients using DIY AID** |  |  |
|  | None | 46 (22.5) |
|  | 1-5 | 77 (37.7) |
|  | 6-14 | 26 (12.7) |
|  | 15-24 | 17 (8.3) |
|  | 25-50 | 4 (2.0) |
|  | >50 | 1 (0.5) |
|  | Unsure | 33 (16.2) |

Respondents were HCP with a variety of practitioner roles; 67 (32.8%) registered dieticians (RD), 65 (31.9%) registered nurses (RN), 58 (28.4%) MD Endocrinologists and 7 (3.5%) MD in other specialties. The majority of practitioners had been working with people with type 1 diabetes for more than six years; 6-10 years, 57 (27.9%), 11-20 years 47 (23%) or greater than 20 years 49 (24%), largely as part of a diabetes clinic or program, 178 (87.3%).

### Current experience and attitudes towards AID

The majority of respondents felt very comfortable in supporting their patients with both CSII (116, 56.9%) and glucose sensor (real time or intermittently scanned CGM) use (150, 73.5%). While most HCP reported feeling comfortable supporting Commercial AID 141 (72.7%), only a minority 42 (21.6%) felt the same for DIY AID, the most frequent response being that participants were not at all comfortable supporting the use of DIY AID systems (64, 33%), figure 1. Comfort levels subdivided according to HCP role (MD Endocrinology, RN, RD and other; comprising other MDs, pharmacists and prefer not to say), specifically towards Commercial and DIY AID systems are shown in figure 2 and table 2. There was no significant difference in technology comfort according to HCP role; CSII kw=3.162, p=0.333, Sensors kw=0.250, p=>0.999, Commercial AID kw=2.279, p=0.467 and DIY AID kw=4.848, p=0.067, although greatest comfort with DIY AID was expressed by RN.

**Figure 1.**
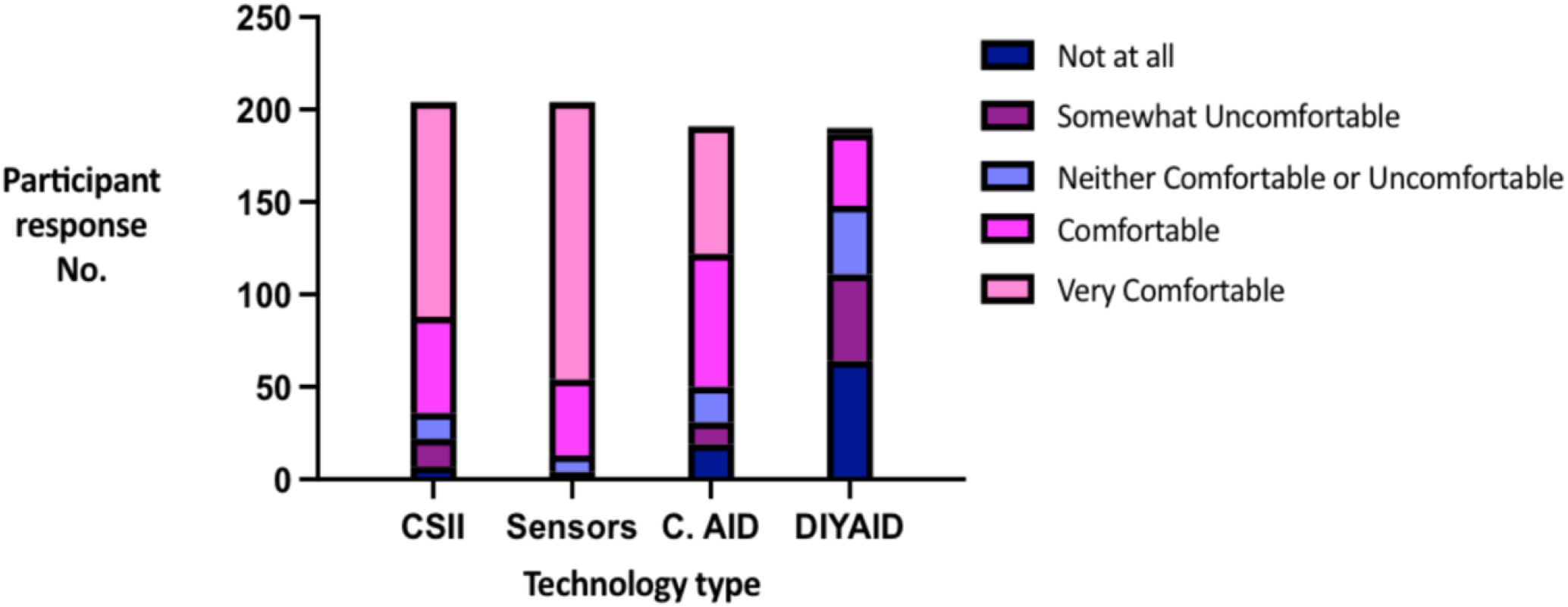
Healthcare Provider current comfort levels in supporting technology use.

**Figure 2.**
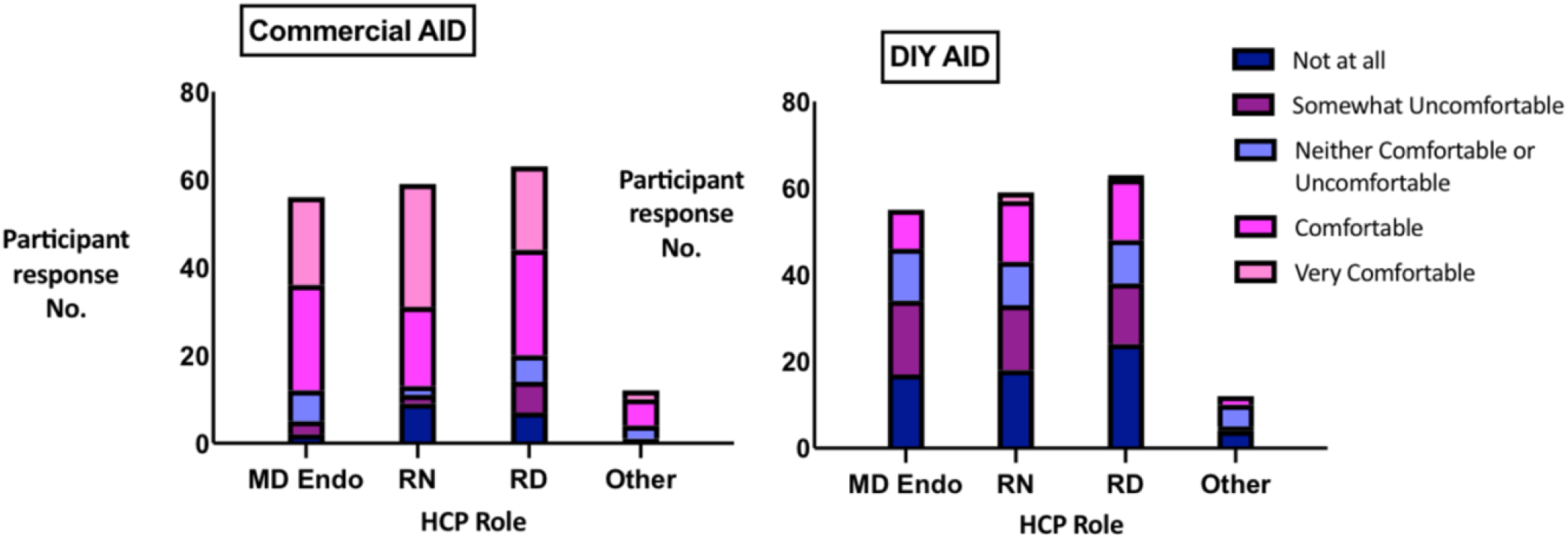
Comfort levels with Commercial and DIY AID systems subdivided according to Healthcare Provider role.

**Table 2.**
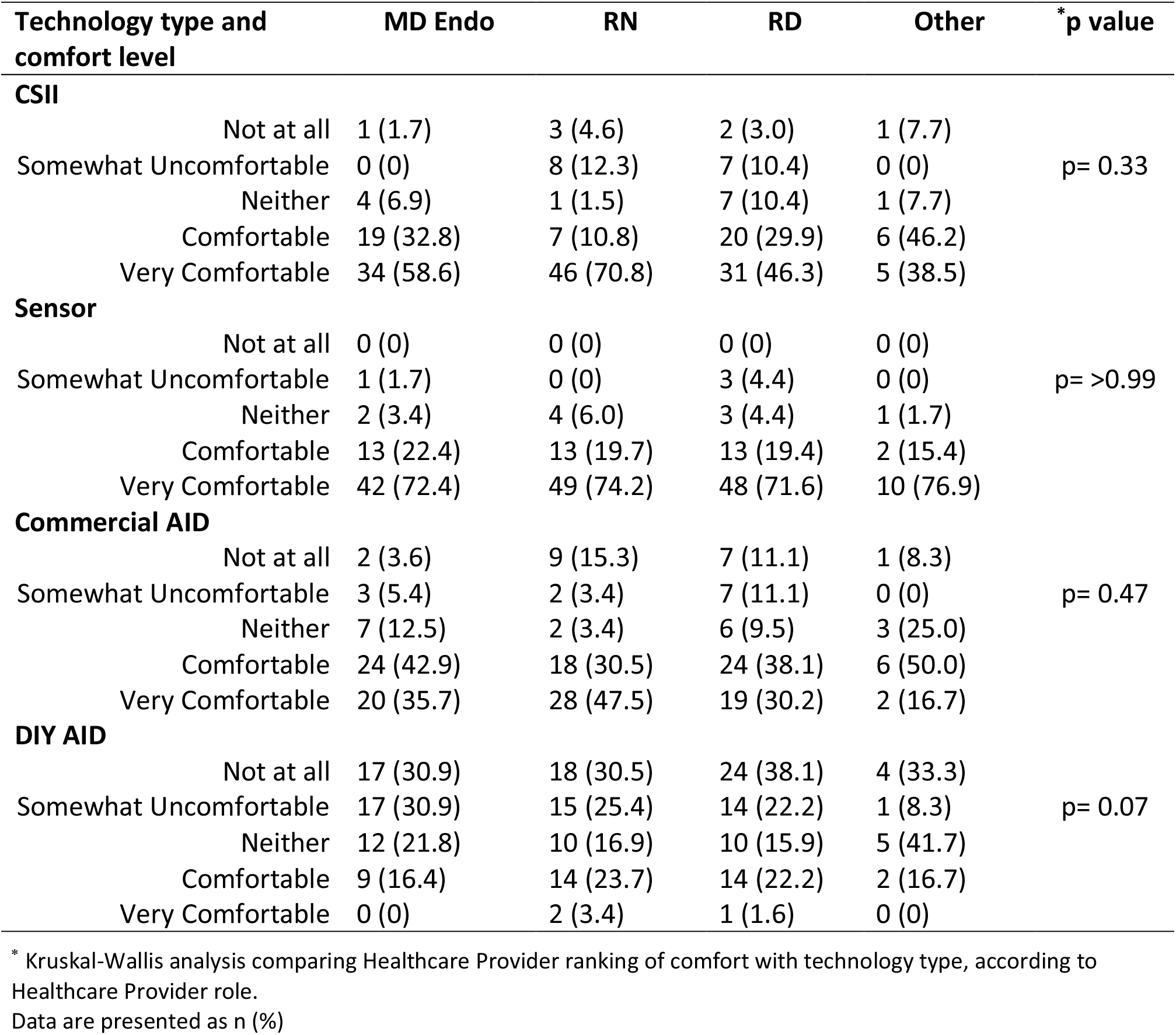
Technology comfort in different Healthcare Provider roles.

The median practice size was 100-500 patients with T1D, with median 25-49% CSII users, 50-75% using sensors, 6-24 patients Commercial and 1-5 patients DIY AID (figure 3). A moderate but significant association was seen between reported comfort levels and proportion of patients in HCP practice using CSII (r=0.5234, p<0.0001), sensors (r=0.2997, p<0.0001), Commercial (r=0.5675, p<0.0001) and DIY AID (r=0.4532, p<0.0001). The number of respondents that expressed feeling comfortable with technologies was greatest for glucose sensors, followed by CSII, and Commercial with least comfort for DIY AID. There was a significant difference in comfort dependent upon device type; kw=9.231, p=0.02, and post hoc analysis revealed this mean rank difference to be greatest between Sensors and DIY AID (p=0.02).

**Figure 3.**
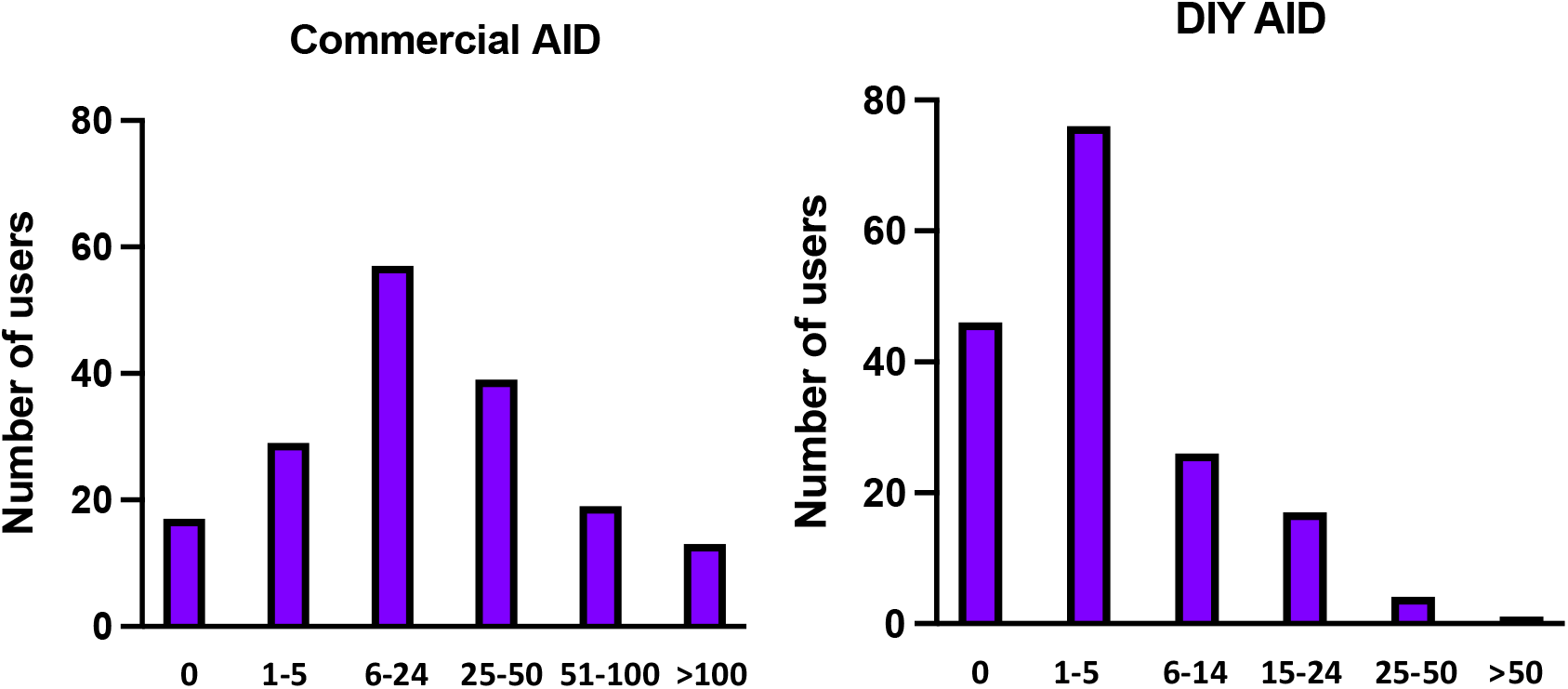
Number of Commercial and DIY AID system users.

With reference to DIY AID systems, HCP most frequently reported that they never initiated discussions with their patients about these systems (94, 48.5%), this was despite 87 (44.8%) of respondents describing themselves as being slightly or much more supportive of DIY AID technology than other diabetes professionals and 106 (60.2%), of respondents advised they would probably or definitely support a patient or family member’s decision to start using a DIY system (table 3).

**Table 3.** DIY AID current practice dependent on Healthcare Provider role.

| Situation | MD Endo | RN | RD | Other | * p value |
| --- | --- | --- | --- | --- | --- |
| Initiate discussion |  |  |  |  |  |
| Never | 27 (48.2) | 29 (49.2) | 30 (48.4) | 7 (58.3) | p=0.95 |
| Rarely | 16 (28.6) | 16 (27.1) | 15 (24.2) | 2 (16.7) |  |
| Sometimes | 8 (14.3) | 13 (22.0) | 13 (21.0) | 3 (25.0) |  |
| Frequently | 3 (5.4) | 1 (1.7) | 3 (4.8) | 0 (0) |  |
| Always | 2 (3.6) | 0 (0) | 1 (1.6) | 0 (0) |  |
| Relative to other HCP |  |  |  |  |  |
| Much less | 3 (5.4) | 4 (6.6) | 7 (10.9) | 0 (0) | p= 0.56 |
| Slightly less | 4 (7.1) | 5 (8.2) | 3 (4.7) | 1 (8.3) |  |
| Same | 23 (41.1) | 17 (27.9) | 16 (25.0) | 6 (50.0) |  |
| Slightly more | 12 (21.4) | 15 (24.6) | 20 (31.3) | 1 (8.3) |  |
| Much more | 10 (17.9) | 16 (26.2) | 12 (18.8) | 1 (8.3) |  |
| Unsure | 4 (7.1) | 4 (6.6) | 6 (9.4) | 3 (25.0) |  |
| Support DIY start |  |  |  |  |  |
| Definitely not | 1 (1.9) | 3 (5.4) | 1 (1.8) | 0 (0) | p= 0.03 |
| Probably not | 9 (17.0) | 5 (8.9) | 3 (5.3) | 3 (33.3) |  |
| Neither | 13 (24.5) | 18 (32.1) | 12 (21.1) | 2 (22.2) |  |
| Probably yes | 25 (47.2) | 15 (26.8) | 22 (38.6) | 3 (33.3) |  |
| Definitely yes | 5 (9.4) | 15 (26.8) | 19 (33.3) | 1 (11.1) |  |

\* Kruskal-Wallis analysis comparing Healthcare Provider ranking of response to situations, according to Healthcare Provider role.
Data are presented as n (%).

If a patient had started using a DIY system the respondents were asked which aspects of care, they would be willing to provide (figure 4), with 22 (14.4%) advising they would not provide ongoing support, referring their patient to another diabetes clinic/provider.

**Figure 4.**
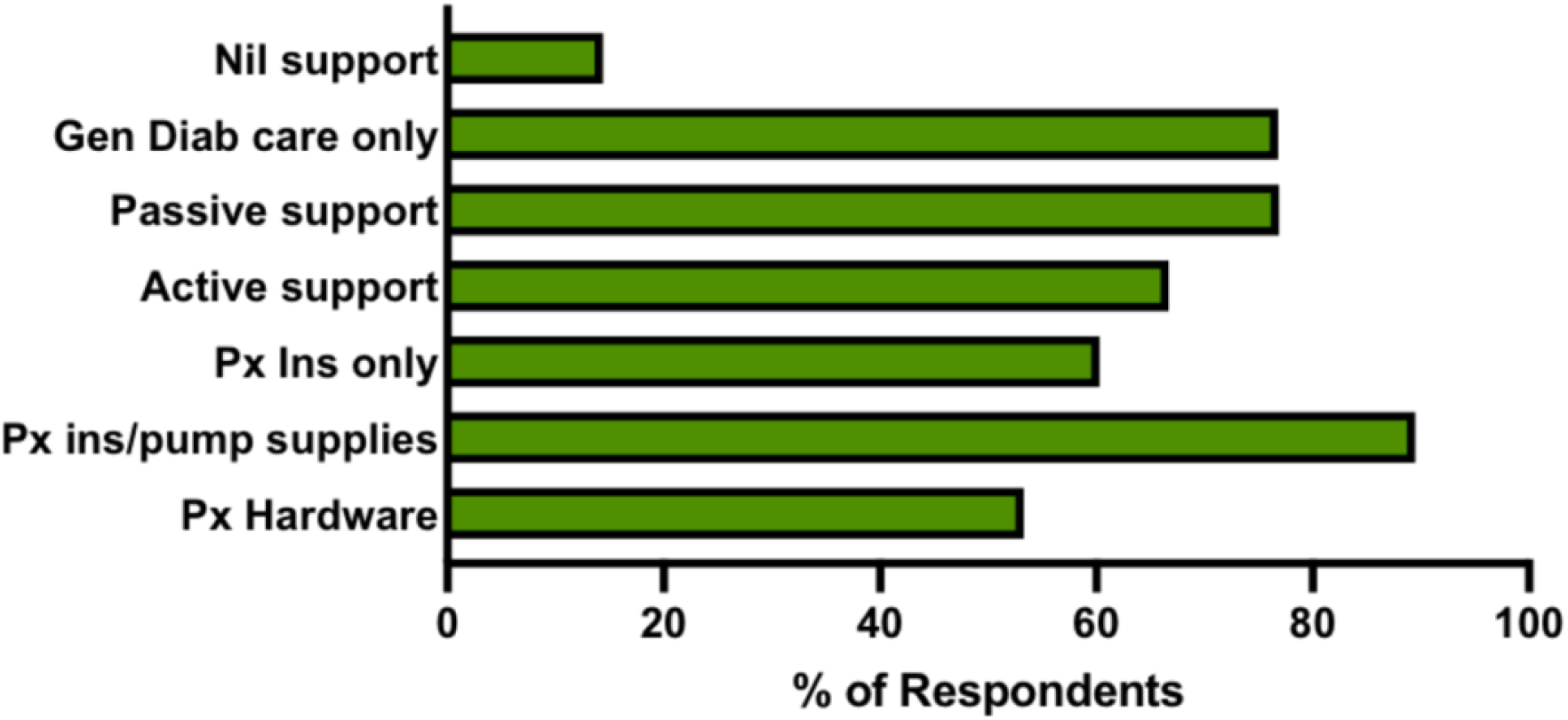
Aspects of care relating to DIY systems that Healthcare Providers are willing to provide.

### Active involvement in DIY AID

Of the 164 participants who responded to this question, 55 (33.5%) stated that they felt themselves to be actively involved in the care of patient’s using DIY systems, these individuals comprised 17 MD Endocrinologists, 24 RN, 12 RD and 2 other HCP (MD other and a pharmacist). These HCP were then asked about the extent of their involvement in reviewing DIY system specific applications, making alterations in settings and discussion of relevant social media platform interactions, in terms of both frequency and comfort (figure 5).

**Figure 5.**
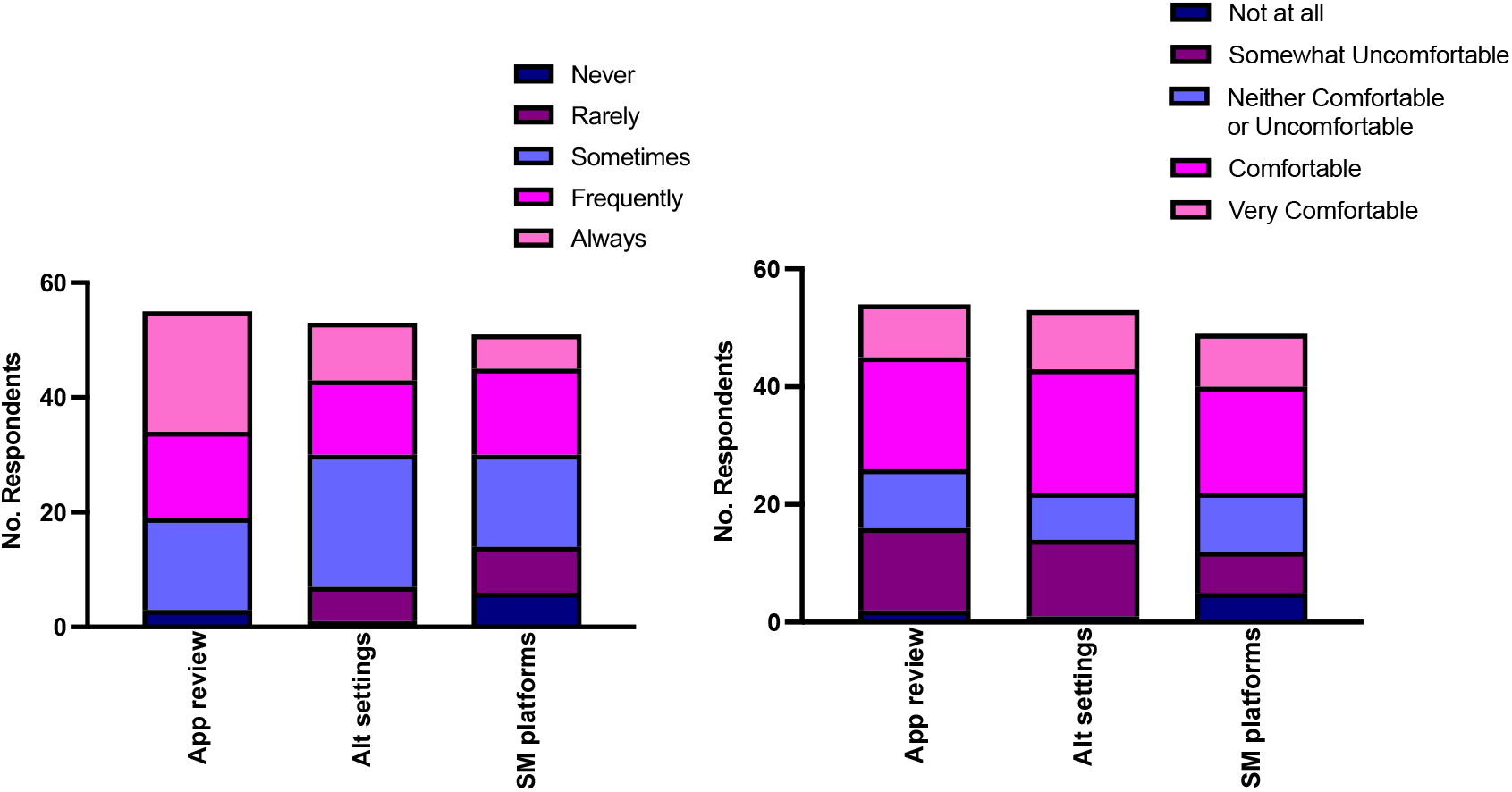
Healthcare Providers deeming themselves to be actively involved in DIY AID ^*^. ^*^ For HCP who deemed themselves to be actively involved in DIY AID use, this graph details response to questions relating to both frequency and comfort with: - App review; HCP review of glucose data through DIY AID specific application eg. Nightscout, Tidepool. - Alt settings; HCP suggesting alteration in DIY AID system settings - SM platforms; HCP discussing with users any specific social media support platforms they are using relating to DIY AID use.

### Barriers to AID use

The perceived barriers which HCP agreed were preventing AID system use are shown in table 4. Funding/coverage for technology was felt to be a barrier in both Commercial; 102(55%) insulin pumps, 151 (81.6%) glucose sensors, and DIY AID systems; 94 (53.1%) insulin pumps, 135 (76.3%) glucose sensors. The greatest perceived barriers to DIY system use were a lack of approved device options (148, 83.6%) and access to staff with system training, 151 (85.3%). Comparison of potential barriers between Commercial and DIY AID systems, revealed a significantly higher number of respondents deemed overall barriers towards DIY relative to Commercial AID (p= 0.001). All proposed barriers were ranked significantly greater for DIY systems relative to Commercial, except funding/coverage for pumps which were equivalent.

**Table 4.** Potential barriers towards AID systems.

| Potential barrier | Commercial AID | DIY AID | * p value |
| --- | --- | --- | --- |
| Funding/coverage for sensors | 151 (81.6) | 135 (76.3) | p=0.0004 |
| Funding/coverage for pumps | 102 (55.1) | 94 (53.1) | p=0.8715 |
| Patient literacy and numeracy | 94 (50.8) | 128 (72.3) | p<0.0001 |
| Patient technological skill | 91 (49.2) | 130 (73.4) | p<0.0001 |
| Few options for officially approved devices | 73 (39.5) | 148 (83.6) | p<0.0001 |
| Lack of access to staff with system training | 60 (32.4) | 151 (85.3) | p<0.0001 |
| Unfamiliar/lack exposure | 33 (17.8) | 130 (73.4) | p<0.0001 |
| Reliability of the system | 32 (17.3) | 74 (41.8) | p<0.0001 |
| Too time consuming | 27 (14.6) | 75 (42.4) | p<0.0001 |
| Lack of high-quality published data | 23 (12.4) | 110 (62.1) | p<0.0001 |
| Medico-legal risks | 18 (9.7) | 114 (64.4) | p<0.0001 |

These perceived barriers to AID use were not significantly different dependent on HCP role (figure 6).

**Figure 6.**
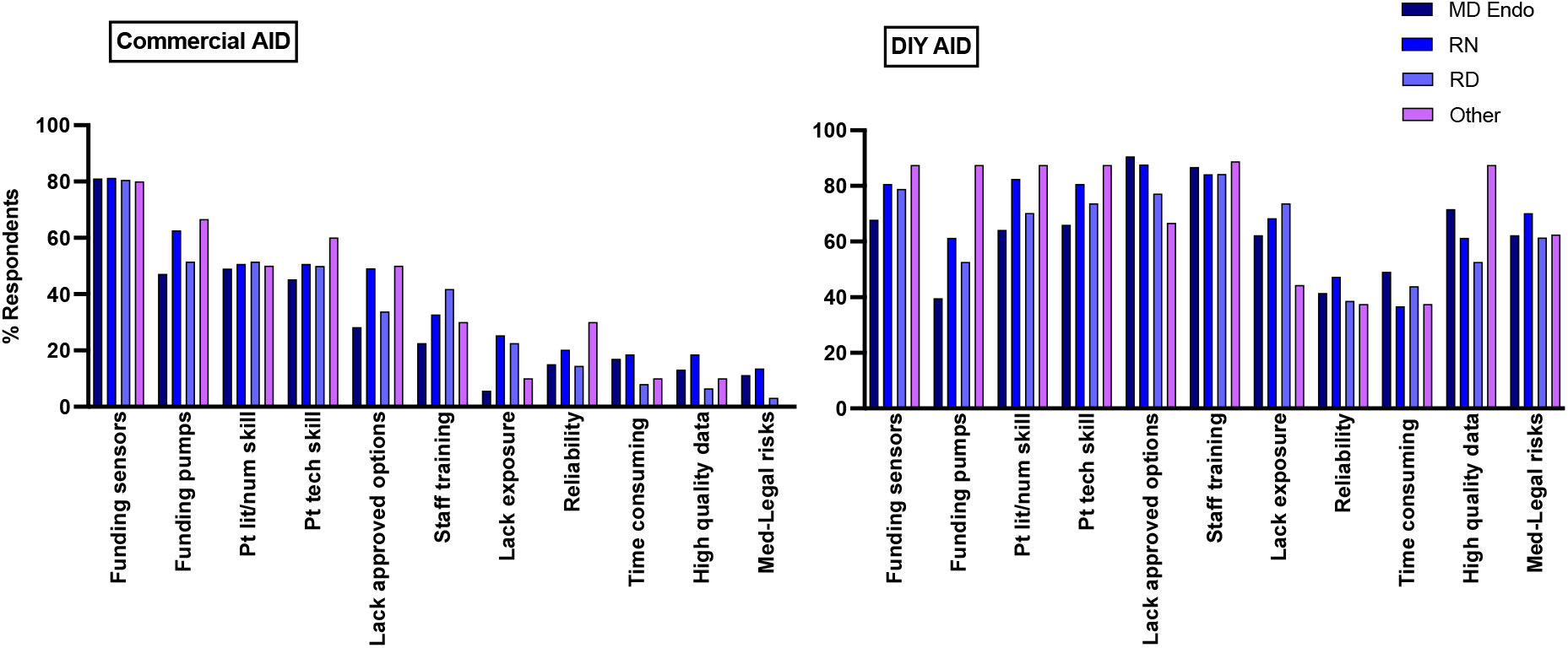
Healthcare Provider perceived barriers to the use of Commercial and DIY AID systems according to Healthcare Provider role.

HCP were asked about characteristics they felt were important for determining suitability of AID (figure 7 and table 5); the most prominent factor identified both for DIY AID systems and Commercial AID was educational level/cognitive ability (155, 91.2% and 152, 89.4% respectively). For all suggested factors, respondents were more likely to deem patients suitable for Commercial AID relative to DIY systems (p=0.004).

**Figure 7.**
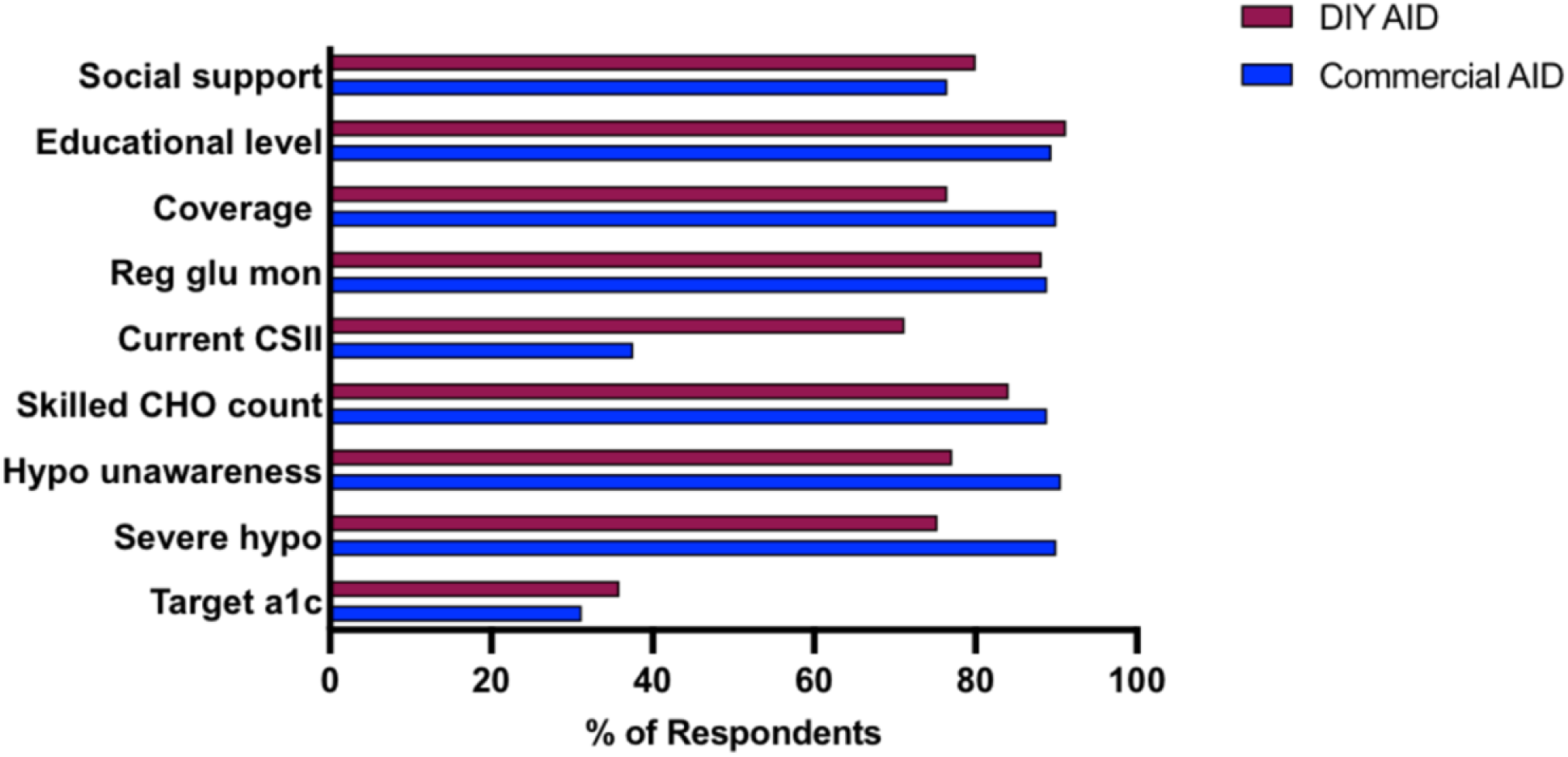
Characteristics deemed to be important by all Healthcare Providers in determining suitability for AID.

**Table 5.** Characteristics important in determining suitability for AID according to Healthcare Provider role.

| Characteristic | MD Endo, n (%) |  | RN, n (%) |  | RD, n (%) |  | Other, n (%) |  |
| --- | --- | --- | --- | --- | --- | --- | --- | --- |
|  | C AID | DIY | C AID | DIY | C AID | DIY | C AID | DIY |
| **HbA1c at/close to target** |  |  |  |  |  |  |  |  |
| Not at all | 12(23.5) | 4(8.2) | 11(21.2) | 8(13.3) | 14(25.0) | 11(19.6) | 2(22.2) | 1(14.3) |
| Not very | 22(43.1) | 16(32.7) | 19(36.5) | 15(25.0) | 27(48.2) | 22(39.3) | 4(44.4) | 2(28.6) |
| Unsure | 4(7.8) | 5(10.2) | 1(1.9) | 7(11.7) | 1(1.8) | 7(12.5) | 0(0) | 3(42.9) |
| Somewhat important | 10(19.6) | 16(32.7) | 16(30.8) | 11(18.3) | 13(23.2) | 12(21.4) | 3(33.3) | 1(14.3) |
| Very important | 3(5.9) | 8(16.3) | 5(9.6) | 19(31.7) | 1(1.8) | 4(7.1) | 0(0) | 0(0) |
| **History Severe Hypo** |  |  |  |  |  |  |  |  |
| Not at all | 0(0) | 1(2.0) | 2(3.8) | 2(4.0) | 2(3.6) | 2(3.6) | 0(0) | 0(0) |
| Not very | 3(5.8) | 3(6.0) | 4(7.7) | 4(8.0) | 4(7.1) | 6(10.7) | 0(0) | 0(0) |
| Unsure | 1(1.9) | 3(6.0) | 0(0) | 2(4.0) | 1(1.8) | 9(16.1) | 0(0) | 3(42.9) |
| Somewhat important | 23(44.2) | 24(48.0) | 16(30.8) | 13(26.0) | 21(37.5) | 19(33.9) | 1(11.1) | 2(28.6) |
| Very important | 25(48.1) | 19(38.0) | 30(57.7) | 29(58.0) | 28(50.0) | 20(35.7) | 8(88.9) | 2(28.6) |
| **Hypo unawareness** |  |  |  |  |  |  |  |  |
| Not at all | 0(0) | 1(2.0) | 2(3.8) | 2(4.0) | 3(5.4) | 2(3.6) | 0(0) | 0(0) |
| Not very | 3(5.8) | 3(6.0) | 3(5.8) | 3(6.0) | 4(7.1) | 6(10.7) | 0(0) | 0(0) |
| Unsure | 1(1.9) | 3(6.0) | 0(0) | 2(4.0) | 0(0) | 7(12.5) | 0(0) | 3(42.9) |
| Somewhat important | 20(38.5) | 20(40.0) | 13(25.0) | 12(24.0) | 18(32.1) | 20(35.7) | 0(0) | 2(28.6) |
| Very important | 28(53.8) | 23(46.0) | 34(65.4) | 31(62.0) | 31(55.4) | 21(37.5) | 9(100) | 2(28.6) |
| **Skilled at CHO counting** |  |  |  |  |  |  |  |  |
| Not at all | 0(0) | 2 (3.9) | 1(1.9) | 0(0) | 0(0) | 0(0) | 0(0) | 0(0) |
| Not very | 4(7.7) | 3(5.9) | 5(9.6) | 3(6.0) | 5(8.9) | 2(3.6) | 0(0) | 0(0) |
| Unsure | 2(3.8) | 2(3.9) | 1(1.9) | 3(6.0) | 1(1.8) | 5(8.9) | 0(0) | 2(28.6) |
| Somewhat important | 23(44.2) | 17(33.3) | 22(42.3) | 18(36.0) | 20(35.7) | 20(35.7) | 5(55.6) | 3(42.9) |
| Very important | 23(44.2) | 27(52.9) | 23(44.2) | 26(52.0) | 30(53.6) | 29(51.8) | 4(44.4) | 2(28.6) |
| **Current CSII use** |  |  |  |  |  |  |  |  |
| Not at all | 6(11.5) | 3(5.9) | 12(23.5) | 5(10.0) | 17(30.4) | 6(10.7) | 1(11.1) | 0(0) |
| Not very | 18(34.6) | 6(11.8) | 17(33.3) | 6(12.0) | 18(32.1) | 4(7.1) | 2(22.2) | 0(0) |
| Unsure | 5(9.6) | 3(5.9) | 3(5.9) | 4(8.0) | 4(7.1) | 4(7.1) | 2(22.2) | 3(42.9) |
| Somewhat important | 12(23.1) | 14(27.5) | 10(19.6) | 11(22.0) | 12(21.4) | 17(30.4) | 2(22.2) | 2(28.6) |
| Very important | 11(21.2) | 25(49.0) | 9(17.6) | 24(48.0) | 5(8.9) | 25(44.6) | 2(22.2) | 2(28.6) |
| **Monitors glucose regularly** |  |  |  |  |  |  |  |  |
|  | 0(0) | 1(2.0) | 0(0) | 1(2.0) | 1(1.8) | 0(0) | 0(0) | 0(0) |
| Not at all | 6(11.5) | 2(3.9) | 5(9.6) | 2(4.0) | 4(7.1) | 1(1.8) | 0(0) | 0(0) |
| Not very | 0(0) | 2(3.9) | 0(0) | 1(2.0) | 3(5.4) | 3(5.4) | 0(0) | 2(28.6) |
| Unsure | 19(36.5) | 15(29.4) | 13(25.0) | 10(20.0) | 17(30.4) | 15(26.8) | 2(22.2) | 3(42.9) |
| Somewhat important | 27(51.9) | 31(60.8) | 34(65.4) | 36(72.0) | 31(55.4) | 37(66.1) | 7(77.8) | 2(28.6) |
| Very important |  |  |  |  |  |  |  |  |
| **Coverage CSII/sensors** |  |  |  |  |  |  |  |  |
| Not at all | 0(0) | 1(2.0) | 1(1.9) | 0(0.0) | 2(3.6) | 1(1.8) | 0(0) | 0(0) |
| Not very | 2(3.8) | 4(8.0) | 5(9.6) | 4(8.0) | 4(7.1) | 7(12.7) | 0(0) | 0(0) |
| Unsure | 0(0) | 4(8.0) | 1(1.9) | 4(8.0) | 1(1.8) | 6(10.9) | 0(0) | 2(28.6) |
| Somewhat important | 19(36.5) | 17(34.0) | 18(34.6) | 19(38.0) | 21(37.5) | 15(27.2) | 4(44.4) | 1(14.3) |
| Very important | 30(57.7) | 24(48.0) | 27(51.9) | 23(46.0) | 28(50.0) | 26(47.2) | 5(55.6) | 4(57.1) |
| **Education/cog. ability** |  |  |  |  |  |  |  |  |
| Not at all | 0(0) | 0(0) | 0(0) | 0(0) | 0(0) | 0(0) | 0(0) | 0(0) |
| Not very | 5(9.6) | 1(2.0) | 3(5.8) | 1(2.0) | 3(5.4) | 0(0) | 1(11.1) | 0(0) |
| Unsure | 3(5.8) | 2(4.0) | 0(0) | 1(2.0) | 3(5.4) | 2(3.6) | 0(0) | 2(28.6) |
| Somewhat important | 20(38.5) | 8(16.0) | 25(48.1) | 15(30.0) | 29(51.8) | 11(19.6) | 4(44.4) | 2(28.6) |
| Very important | 24(46.2) | 39(78.0) | 24(46.2) | 33(66.0) | 21(37.5) | 43(76.8) | 4(44.4) | 3(42.9) |
| **Family resources/support** |  |  |  |  |  |  |  |  |
| Not at all | 0(0) | 0(0) | 1(1.9) | 1(2.0) | 0(0) | 0(0) | 0(0) | 0(0) |
| Not very | 9(17.3) | 4(8.0) | 4(7.7) | 1(2.0) | 5(8.9) | 2(3.6) | 2(22.2) | 0(0) |
| Unsure | 5(9.6) | 4(8.0) | 3(5.8) | 3(6.0) | 11(19.6) | 8(14.5) | 0(0) | 4(57.1) |
| Somewhat important | 21(40.4) | 14(28.0) | 28(53.8) | 21(42.0) | 29(51.8) | 22(40.0) | 3(33.3) | 2(28.6) |
| Very important | 17(32.7) | 28(56.0) | 16(30.8) | 24(48.0) | 11(19.6) | 23(41.8) | 4(44.4) | 1(14.3) |

Participants were asked to note any other prerequisites they felt were essential for AID use (these are described in the word cloud in figure 8), with ability, understanding, expectations, motivation, technology and access the most frequent words used in response to this question.

**Figure 8.**
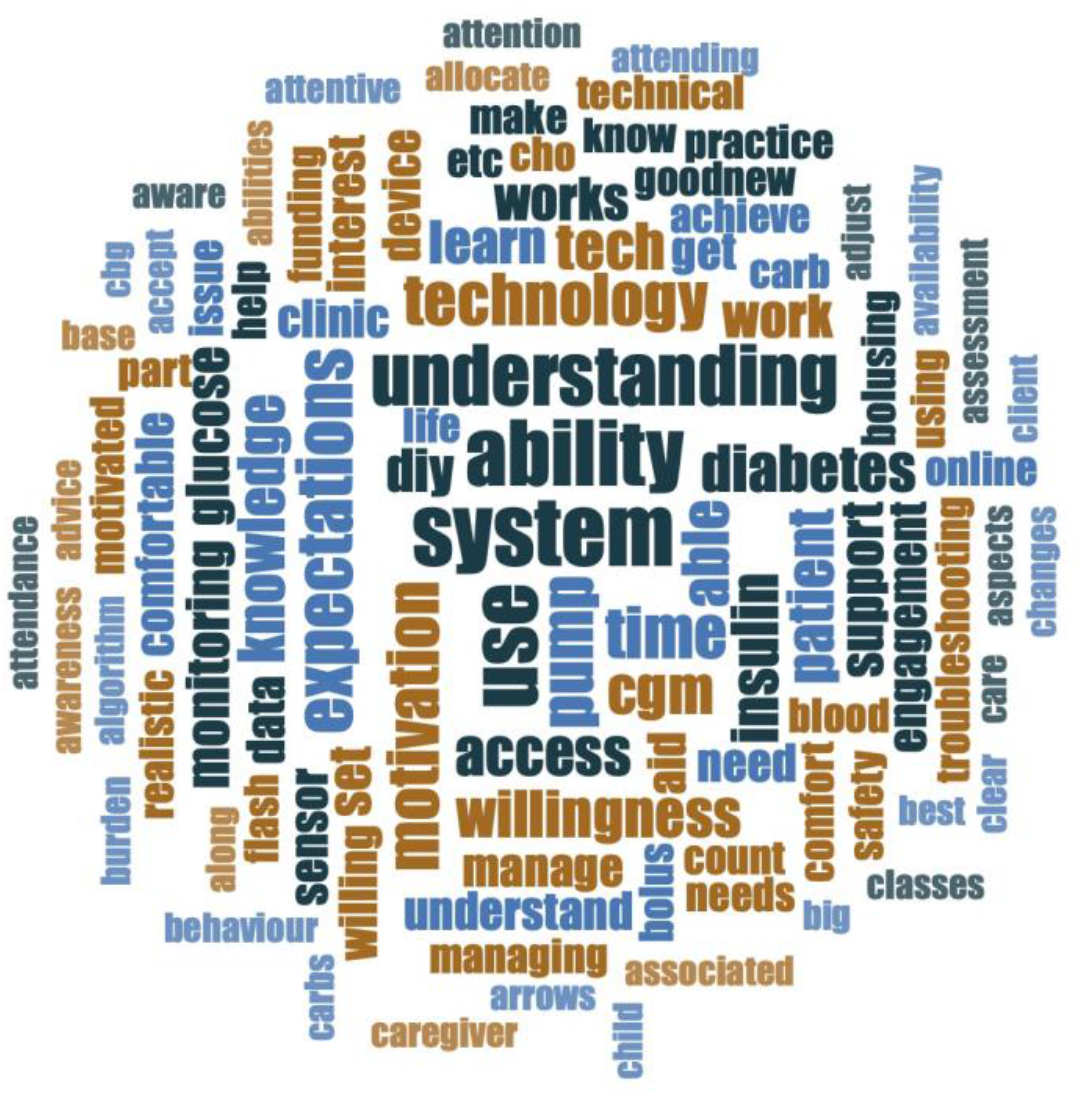
Essential prerequisites for AID use.

HCP were asked if they were aware that any of their patients had stopped using any form of AID; 79 (44.6%) answered that they were aware of someone who had stopped using an AID system. The prominent reasons for stopping AID described by HCP (figure 9) were sensor issues and frustration with the system and its alarms, particularly relating to the Medtronic sensor. The words ‘coverage’ and ‘cost’ appeared frequently in HCP responses to this question.

**Figure 9.**
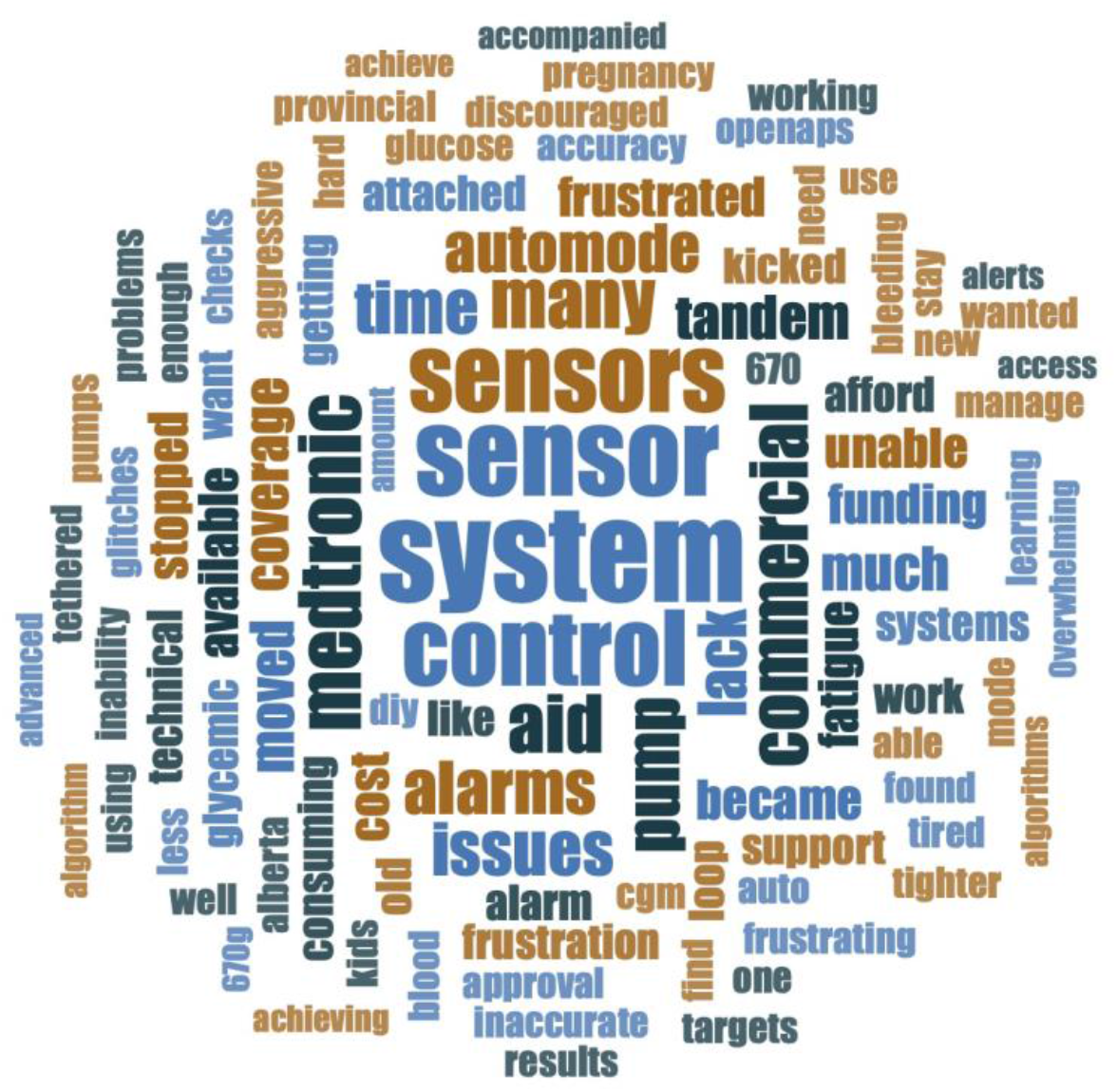
Reasons for users stopping AID.

### AID Scenarios

HCP were asked about their comfort levels with Commercial or DIY AID use in the same proposed patient scenarios (figure 10). In both system types the greatest level of concern was most frequently expressed by respondents in those users ‘infrequently monitoring their glucose levels’; 47.3% Commercial and 60.7% DIY AID, as well as those in the scenario ‘not using the bolus calculator with no set insulin:carbohydrate ratio or insulin sensitivity factor’; 46.1% Commercial and 55.8% DIY AID. Conversely, HCP were most comfortable in the use of these systems in the setting of an individual with a ‘close to target HbA1c’, although comfort with Commercial systems was greater than DIY; 62.3% Commercial and 39.3% DIY AID (p= 0.001).

**Figure 10.**
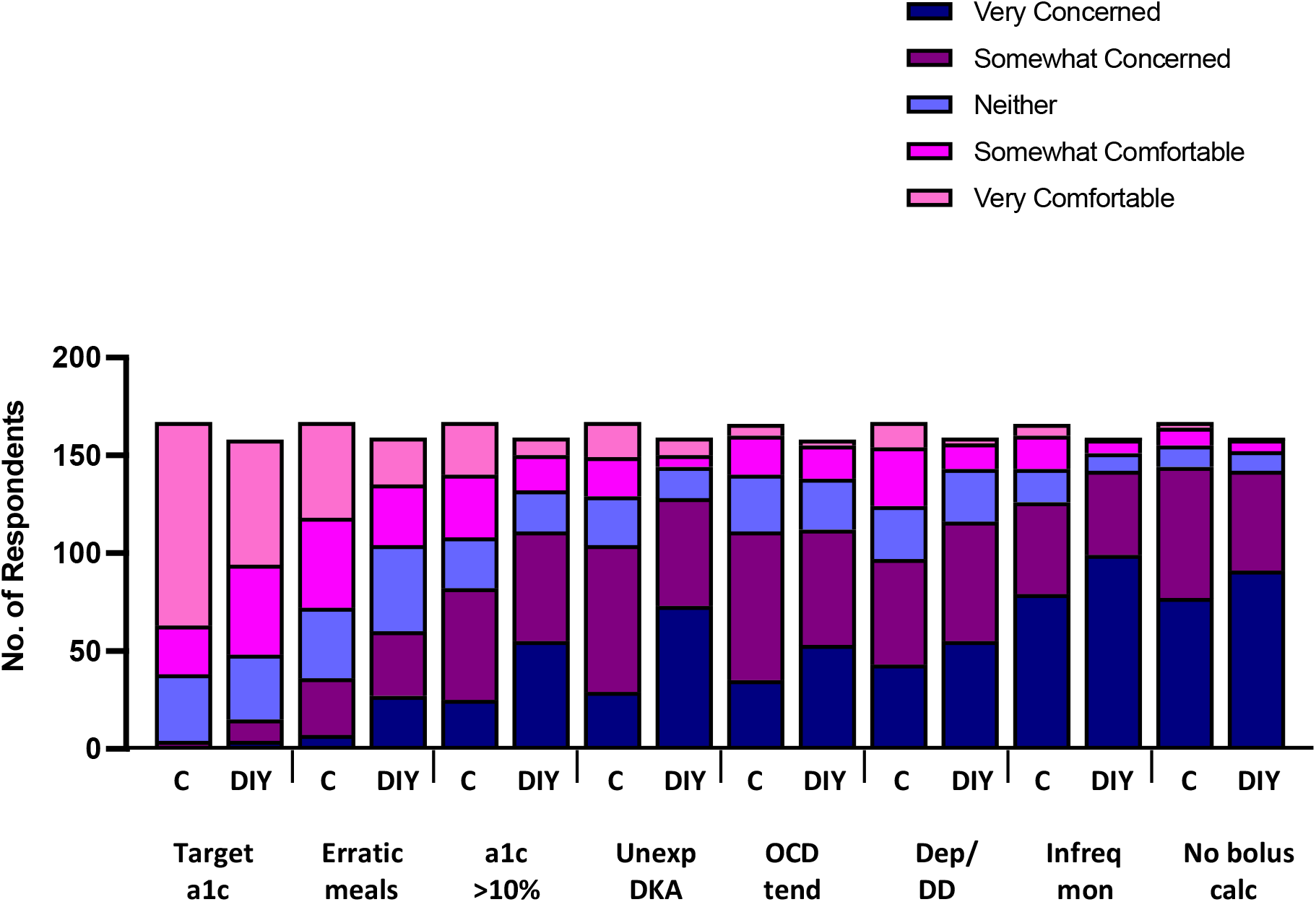
Healthcare Provider comfort in commencing Commercial and DIY AID systems in specific scenarios. ^*^. ^*^ Scenarios posed to HCP with each AID system type; An individual with at or close to target HbA1c, an individual eating erratic meals, an individual with HbA1c consistently >10%, an individual with recent unexplained DKA (in the preceding 12 months), an individual with OCD tendencies, an individual with evidence of Depression or Diabetes-related Distress, an individual who is infrequently monitoring their blood glucose levels and an individual who is not using the bolus calculator, with no set insulin:carbohydrate ratio or insulin sensitivity factor.

Comfort levels, specifically of MD Endocrinologists were reviewed for both Commercial (n=51) and DIY AID systems (n=50). With Commercial systems, this group of HCPs were very concerned with the use of these systems in those individuals ‘infrequent glucose monitoring’ (20, 39.2%) and ‘not using bolus calculator’ (21, 41.2%). Concern with DIY AID use was expressed in individuals with an ‘HbA1c consistently >10%’ (27, 54%) and ‘an episode of unexplained DKA in the preceding 12 months’ (25, 50%), in addition to ‘infrequent glucose monitoring’ (35, 70%) and ‘not using bolus calculator’ (28, 56%).

### Potential Enablers of AID

When asked about potential interventions to improve HCP confidence in recommending either a Commercial or DIY AID system, at least two thirds of participants (113, 66%) answered positively to each suggested option. User support (147, 90.7%) and user education (145, 89.6%) were the most popular responses relating to Commercial AID, and user education (154, 95.7%) and HCP education (153, 95.1%) for DIY AID systems. MD Endocrinologists were the most frequent respondents suggesting the need for implementation of medico-legal guidance relating to DIY AID systems, 93.9% of Endocrinologists felt this was required (figure 11). Significantly more respondents deemed that the suggested potential interventions were required to improve HCP confidence in recommending DIY, relative to Commercial AID systems, p=0.0005. No significant difference was seen in suggested intervention according to HCP role; Commercial (KW=0.265, p=0.876) and DIY systems (KW= 0.110, p=0.946).

**Figure 11.**
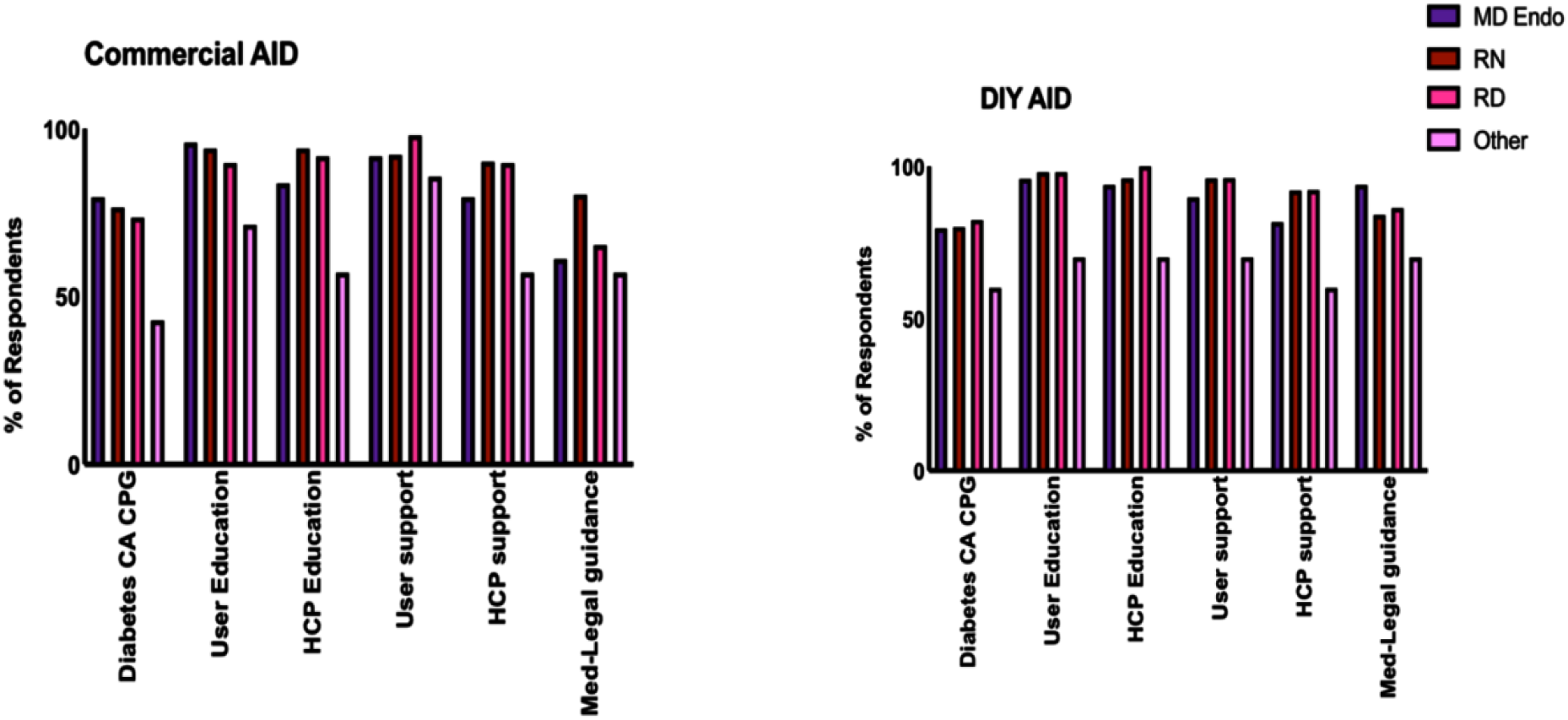
Potential interventions felt appropriate to improve AID confidence for different Healthcare Provider roles.

## Discussion

This cross-sectional study is the first to examine perspectives on the current use of AID systems (Commercial and DIY), from multidisciplinary HCP caring for adults and children with T1D from across Canada. In this survey of HCP with large type 1 diabetes clinics, with a high proportion of technology use in the form of both insulin pumps and glucose sensors, a low number of users of AID systems were reported. There were a greater number of users of Commercial relative to DIY AID, with greater HCP comfort in supporting Commercial AID use. HCP reported similar pre-requisites and cautions for safely initiating the technology with both AID types. Education, for both HCP and users, were identified as areas of intervention to increase HCP confidence in recommending AID.

Similar user numbers of DIY systems were reported by Canadian HCP to those in the 2019 UK HCP survey; 85% reported 0-5 users [12]. These figures may be imprecise, dependent on memory and recall from participants. Additionally, specifically relating to DIY AID use, HCP may not always be aware that their patients are using these systems. This may be something that a user would currently worry about disclosing to their HCP, due to concerns regarding potential technology removal or discharge from a particular clinic or provider’s care [13]. Although difficult to ascertain precise figures, it is estimated that there are now over 10,000 users of DIY AID systems worldwide [14]. User survey data suggests Europe (notably Germany and the UK), North America, Australia and South Korea to be the most prominent locations, but an exact figure of the number of Canadian users of DIY AID systems is not currently available [15].

There was greater comfort with Commercial than DIY systems, although almost half of respondents deemed themselves to be more supportive of DIY systems than other HCP colleagues. Despite this, few HCP would initiate discussions relating to DIY AID and more would provide permissive support with ongoing prescription of component devices. Some HCP (14.4%), did express that they would withdraw care of patients using a DIY system, confirming patient fears of disclosing DIY AID use. Similarly, in the UK HCP survey respondents; a high proportion expressed that they would not initiate conversations with their patients about DIY AID systems (91%), but were willing to support users (55%), and most would continue to provide ongoing care (94%) [12]. The positive responses gathered relating to DIY comfort and support are not in line with the views of multi-national users of DIY AID systems; data suggests that the majority of DIY AID users, do not feel HCP in general have a good understanding of these systems [8]. With the self-selecting nature of responding to a survey, there is the potential for bias in the responses, with the prospect of this sample being from a skewed HCP population viewpoint, relating to technology experience and comfort. These user experiences may reflect diabetes care teams with less technology experience and involvement.

Cost was an important factor raised by participants resulting in restricted access to both of these systems, with insulin pumps and CGM resulting in significant financial outgoings for the user if they do not have funding or coverage for these devices. This is estimated to be $6000-7000 CAD for an insulin pump, $3000 CAD for yearly pump supplies and $3000-$6000 CAD annually for real time Continuous Glucose Monitoring (rtCGM) [16, 17]. Although more expensive than alternate forms of insulin delivery, the use of Commercial AID is cost-effective [18]. However, as a result of provincial funding models in Canada, access remains unequal, with insulin pump use more common in areas with reimbursement programs in place [19]. DIY AID system users, in addition to an insulin pump and rtCGM, have the added costs of ensuring a suitable phone or watch interface, a communicating device or microcomputer, the subscription for a developer’s license to build the relevant application as well as an appropriate computer platform to build it on [20]. Unfortunately, these described overwhelming costs to the user are likely to continue to be an ongoing barrier to the broader use of AID, unless significant changes to coverage for diabetes technologies occurs, to improve uniformity of access across Canada.

Having an at or close to target HbA1c was identified to be an optimal scenario in which to commence AID. Studies in the use of both Commercial and DIY AID systems have highlighted improvements in glycemic outcomes, demonstrated by both time in range (TIR) and HbA1c level [21, 22]. There is the potential for these systems to improve glucose control and reduce hypoglycemia [23]. Diabetes Canada recommends using Commercial AID to improve or maintain HbA1c, without increasing hypoglycemia, especially in individuals experiencing nocturnal hypoglycemia [24]. Similarly in the Diabetes UK technology pathway, Commercial AID systems are recommended in individuals with an HbA1c remaining above 8.5%, despite a single form of technology use (CSII or CGM) [25]. Our survey responses bring into question whether access to Commercial AID systems may be restricted unnecessarily, relating to an individual’s current glycemic outcomes whilst using an alternative method of insulin delivery. HCP comfort may be contributing to inequitable care, further exacerbating the existing barriers to AID as a result of financial costs and coverage.

Concern was highlighted by HCP in initiating either type of AID system if an individual is infrequently monitoring their glucose levels. To their HCP this may prompt concern about lack of engagement in diabetes management and potentially treatment compliance [26]. Each of these AID system types incorporate rtCGM in combination with an insulin pump, with rtCGM enabling automated recording of glucose levels, irrespective of frequent user input or action. Implementation of CGM use alone, without CSII or AID, is associated with improved glycemic control and a reduction in frequency of hypoglycaemia [27, 28]. While active self-management is required to mitigate risks (eg of DKA in the case of infusion set malfunction), denying access to a system that requires less user-input because of infrequent glucose monitoring appears counterintuitive.

A diagnosis of type 1 diabetes brings with it a vast amount of new information, and educational needs, often at a young age. To aid this process, structured education is key, in addition to contact with, and support from HCP, in supporting the person with diabetes with this diagnosis and its implications on their day-to-day life [25, 29]. Carbohydrate counting; to enable flexible dietary intake with optimal matched insulin delivery, the development of individual insulin to carbohydrate ratios (ICR) and insulin sensitivity factors (ISF), are crucial in optimizing glycemic outcomes, whilst utilizing multiple daily injections of insulin or traditional pump therapy [30]. HCP responding to this survey were reluctant to commence AID in the setting of ‘an individual not using a bolus calculator, no set ICR or ISF’. A lack of understanding around, and implementation of, these mathematical settings, reflect a likely deficiency in diabetes-related education and crucial knowledge acquisition [31]. However, due to the nature of the algorithm incorporated in AID systems, it may be argued that this concept is less important, with the system allowing flexibility to overcome inaccuracies in carbohydrate counting. With these automated capabilities it could be considered possible for HCP themselves to program the settings, initiate and continue AID system use in a person with minimal diabetes-related education. This concept is not optimal, heavily relying upon the technology functioning as described, any system failure in this setting could result in significant harm for the user, but this may not be any greater than the risks of conventional pump failure or infusion site issues.

HCP with greater experience in the use of AID systems reported greater comfort in supporting their use. Rapid advancements, especially demonstrated in the development of user-driven DIY systems, have resulted in minimal HCP experience and low comfort levels. Unlike the Commercial systems, with no device or pharmaceutical company at the forefront of this progression, no specific targeted education for HCP has been produced in the use of DIY systems. HCP are left in the position of noticeable knowledge gaps in the understanding of DIY AID systems, which their patients may have now implemented as their chosen glucose management system, these knowledge deficiencies were reflected in the American Association of Diabetes Educators survey [8]. There additionally remains ongoing ethical and legal uncertainty for HCP, in supporting patients who are using DIY AID systems, due to the unregulated and unapproved nature of these devices. There is a lack of consensus specialist guidance available [32]. Attitudes towards DIY AID collected were generally similar across HCP disciplines, although medico-legal concerns were most prominent among physicians.

## Conclusion

The results of this survey have given a snapshot view of the current approach and practices of HCP throughout Canada, towards the use of AID systems. Limitations in AID availability due to funding or coverage of technology are apparent. These data suggest that training of providers and recommendations around best practice would be helpful for practitioners, and may also clarify medico-legal uncertainty.

Further data collection will be beneficial, including expansion of the scope of this survey to incorporate responses from more HCP, in different countries worldwide. This will enable a greater variety of experiences to be gathered, and a greater understanding of potential and beneficial interventions to be developed. The perspectives from AID system users, especially relating to their healthcare experiences, are also imperative to understand. Incorporation of user, and broader HCP knowledge and perspectives, will hopefully result in improved access to the benefits of AID use for more people with type 1 diabetes.

## Data Availability

All data produced in the present work are contained in the manuscript

