## appendices for "Canadian Healthcare Providers’ Attitudes Towards Automated Insulin Delivery Systems"

Thank you for taking this 20-minute, anonymous survey. Your name will not be recorded anywhere in this survey. You may choose not to answer any questions you don’t want to answer, by selecting the option prefer not to say or n/a.

About the researchers: This survey is part of a study led by Dr Anna Lam and Dr Peter Senior, both endocrinologists and researchers at the University of Alberta, specializing in type 1 diabetes management and currently looking after patients with type 1 diabetes using Automated Insulin Delivery systems (AID). Helping us with the project are Dr Holly Witteman and Olivia Drescher, researchers from Laval University; and a patient partner from Diabetes Action Canada, Kate Farnsworth, who runs the Looped Facebook group.

We are also collaborating with colleagues in the UK; Dr Emma Wilmot and Dr Tom Crabtree, who conducted a brief HCP survey in 2019 (Crabtree T, Choudhary P et al. Health-care Professional Opinions of DIY Artificial Pancreas Systems in the UK. Lancet Diabetes and Endocrinology.2020;8(3):186-187).

Purpose of the study: The purpose of this study is to understand the current knowledge, experience and attitudes of healthcare providers across Canada in the use of Automated Insulin Delivery systems (AID) in patients with type 1 diabetes. Questions relate to the use of Commercial AID; Hybrid Closed Loop systems (Medronic 670G or 770G and Tandem Control IQ), as well as DIY AID systems; AndroidAPS, OpenAPS and Loop. We aim to identify areas of knowledge gaps and concern, to enable targeted education with the aim of improved patient care for those using these systems.

Your participation: Participating in this study will involve answering a 20-minute survey, with 31 questions, on your current experience, knowledge and attitudes towards AID systems.

Eligibility:

You may participate in this study if:

You are a healthcare provider licensed in CanadaYou look after patients with type 1 diabetes as part of your professional role; including physicians, nurses, dietitians and educatorsYou are able to read and understand this pageRisks and benefits: There are no personal risks or benefits to answering the survey. Your answers may help guide future education in the area of AID systems and therefore improved patient care for people living with type 1 diabetes and using these systems.

Confidentiality and data protection: All information we collect will be confidential and used only for research purposes. Data will be collected and stored on secure servers located in Canada, it will be anonymous, with no identifiable information. When we work with data, the only people who will have access to the data will be the investigators in our research team, who have completed relevant ethics training. When the data is stored on our computers, each computer will be secure and password protected.

After the end of the study: Only anonymous, non identifiable data will be stored. Data will be kept securely, in line with the University of Alberta Ethics regulations, for a minimum of five years.

Contacts: If you have any questions about the research study, or if you experience a problem as a result of participating in the study, please contact:

Dr Anna Lam

Department of Medicine, University of Alberta

Consent: By answering the questions in this survey, you are indicating your consent to have your answers used in this study. If you do not want to participate in the study, please do not answer the questions.

This study has been approved by the Research Ethics Committee of University of Alberta, Research Ethics ID; Pro00108472.

**Healthcare provider and practice characteristics**

1. Are you a healthcare provider licensed to practice	Yes

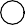

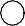

in Canada?	No

Thank-you for your interest but this survey is restricted to HCP licensed to practice in Canada

2. Do you provide care for adults or children with	Adults

type 1 diabetes?	Children

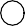

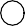

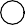

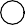

Both Neither

Thank-you for your interest but this survey is restricted to HCPs looking after people with type 1 diabetes

3. Where do you practice?	Alberta

British Columbia Manitoba

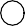

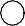

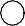

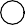

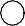

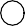

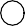

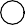

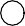

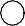

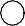

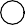

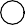

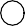

New Brunswick Newfoundland and Labrador Northwest Territories

Nova Scotia Nunavut Ontario

Prince Edward Island Quebec Saskatchewan Yukon

Prefer not to say

4. Which setting best describes your clinical	Academic centre

practice?	Urban hospital

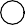

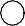

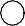

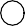

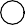

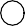

Rural hospital Community- urban Community - rural Prefer not to say

5. What kind of practitioner are you?	MD Endocrinology MD Internal Medicine MD Family Medicine MD other

RN RD

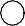

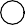

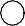

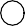

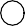

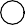

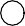

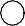

Pharmacist Prefer not to say

6. Are you a designated CDE?	Yes

No

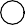

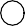

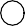

Prefer not to say

7. How long have you worked with people with diabetes	< 1 for? (years)	1-5

6-10

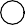

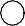

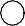

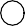

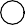

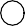

11-20

>20

Prefer not to say

8. a) Do you practice individually or as part of a diabetes clinic/program?

Individually

Part of diabetes clinic/program Prefer not to say

8. b) Questions 9-11 and 14-15 will relate to patient	Individual practice numbers in your current practice, do you wish to	Diabetes clinic/program

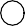

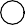

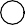

answer these based on your individual practice or	Prefer not to say those of your diabetes clinic/program?

9. Roughly, how many of your patients have type 1	< 10

diabetes?	10-50

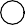

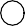

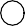

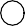

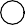

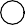

51-100

101-500

>500

Don't know

10. What proportion of your patients with type 1	< 5%

diabetes use insulin pumps?	5-24%

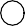

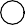

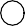

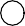

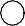

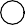

25-49%

50-75%

>75%

Don't know

11. What proportion of your patients with type 1	< 5%

diabetes use Flash (Freestyle Libre) or Continuous	5-24%

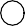

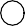

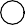

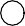

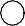

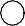

Glucose Monitoring (CGM)?	25-49%

50-75%

>75%

Don't know

12. How comfortable do you feel supporting your	not at all

patients that are using insulin pumps?	somewhat uncomfortable

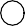

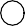

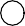

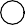

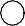

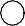

neither comfortable or uncomfortable comfortable

very comfortable n/a

13. How comfortable do you feel supporting your	not at all

patients that are using Flash (Freestyle Libre) or	somewhat uncomfortable

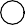

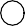

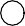

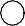

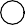

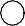

Continuous Glucose Monitors (CGM)?	neither comfortable or uncomfortable comfortable

very comfortable n/a

**Healthcare provider current experience and attitudes towards Automated Insulin Delivery**

**(AID)**

14. How many of your patients are using Commercial AID	None systems (Hybrid Closed Loop such as Medtronic 670G,	1-5

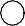

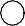

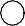

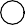

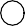

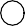

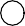

Tandem Control IQ)	6-24

25-50

51-100

>100

Don't know

15. How many of your patients, that you are aware of,	None are using Do-it-Yourself (DIY) AID systems (eg Loop,	1-5

Open APS, Android APS)?	6-14

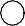

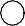

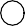

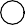

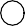

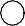

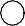

15-24

25-50

>50

Don't know

16. How comfortable do you feel supporting your	not at all

patients with Commercial AID systems?	somewhat uncomfortable

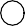

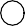

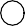

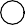

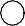

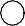

neither comfortable or uncomfortable comfortable

very comfortable n/a

17. How comfortable do you feel supporting your	not at all

patients with DIY AID systems?	somewhat uncomfortable

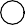

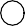

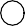

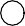

neither comfortable or uncomfortable comfortable

very comfortable n/a

18. Do you initiate discussions about DIY AID as a	never

treatment option in your consultations?	rarely sometimes frequently always

n/a

19. Compared with other diabetes professionals, how	much less supportive of DIY AID technology do you think you are?	slightly less

about the same slightly more much more unsure

**Barriers to AID use**

20. For a patient considering Commercial or DIY AID	Commercial

systems, which are you more likely to recommend?	DIY Either Neither Unsure N/A Other

If recommending other system, please highlight what this would be?

**20 a). For COMMERCIAL AID systems, to what extent do you agree that the following act as barriers for you recommending these systems?**

**Please rank these factors strongly disagree to strongly agree, or n/a).**

You are unfamiliar with/lack exposure to AID systems

Too time consuming

Few options for officially approved devices

Reliability of the system Medico-legal risks

Lack of high quality published data

Patient technological skill Patient literacy and numeracy Funding/coverage for pumps Funding/coverage for sensors

Lack of access to staff with AID training

strongly

disagree

disagree	neutral	agree	strongly agree	n/a

**20 b). For DIY AID systems to what extent do you agree that the following act as barriers for you recommending these systems?**

**Please rank these factors using options strongly disagree to strongly agree, or n/a)**

You are unfamiliar with/ lack exposure to DIY AID systems

Too time consuming

Lack of official approval of devices/systems

Reliability of the system Medico-legal risks

Lack of high quality published data

Patient technological skill Patient literacy and numeracy Funding/coverage for pumps Funding/coverage for sensors

Lack of access to staff with DIY AID training

strongly

disagree

disagree	neutral	agree	strongly agree	n/a

21. Are you aware of any of your patients having	Yes

stopped using AID systems?	No

Unsure

21. a) If so, what reasons did they give for stopping?

**AID Scenarios**

**In some of the following scenarios (Q24/25, Q26/27, Q30/31) we will ask you to firstly answer relating to COMMERCIAL AID systems (eg Medtronic 670G, Tandem Control IQ) and then relating to DIY AID systems (OpenAPS, AndroidAPS and Loop).**

22. If a patient or patient family member asked you	definitely not

whether you would support their decision to START	probably not

using a DIY AID system, to what extent would you	neither encourage or discourage support/encourage them?	possibly yes

definitely yes n/a

**23. If a patient (or family) HAS STARTED using DIY AID on their own, which aspects of the following support would you provide them with?**

**Please rank from definitely not to yes definitely, or n/a.**

definitely not	probably not	unsure	yes possibly	yes definitely	n/a

Facilitate Prescriptions for	hardware (pumps/sensors)

Facilitate Prescriptions for insulin	and pump supplies

Facilitate Prescriptions for insulin	only

Active support - help with pump	settings and adjustments

Passive support- direct to	online/other resources

On-going support only for	general diabetes care

No on-going support - refer to	another diabetes clinic/provider

**24. When considering COMMERCIAL AID systems, how important do you think the following**

**factors are in determining whether you would consider a patient 'suitable' for these systems? Please rank from not at all to very important, or n/a.**

HbA1c at or close to target History of severe hypoglycemia Hypoglycemia unawareness Skilled at carbohydrate counting Current pump user

Monitors glucose levels regularly and reliably

Has coverage/funding for pumps and sensors

Educational level/cognitive abilities

Family resources/social support

not at all	not very	unsure	somewhat

important

very

important

n/a

**25. When considering DIY AID systems, how important do you think the following factors are in determining whether you would consider a patient 'suitable' for DIY AID?**

**Please rank from not at all to very important, or n/a.**

HbA1c at or close to target History of severe hypoglycemia Hypoglycemia unawareness Skilled at carbohydrate counting Current pump user

Monitors glucose levels regularly and reliably

Has coverage/funding for pumps and sensors

Educational level/cognitive abilities

Family resources/social support

not at all	not very	unsure	somewhat

important

very

important

n/a

**26. When starting a COMMERCIAL AID system, how comfortable or concerned would you be about an individual starting Commercial AID in the following scenarios?**

**Please rank very concerned to very comfortable, or n/a.**

very

concerned

slightly

concerned

neither

concerned or comfortable

somewhat

comfortable

very

comfortable

n/a

Unexplained episode of DKA in	the last 12 months

HbA1c at or close to target in	current pump user

HbA1c consistently >10%	Infrequent glucose monitoring	Erratic schedule/meal pattern

Evidence of depression and/or	diabetes related distress

Obsessive compulsive	tNeontduesnincigesbolus calculator - no

set insulin: carb, or insulin sensitivity factor

**27. When commencing a DIY AID system, how comfortable or concerned would you be about an individual starting DIY AID in the following scenarios?**

**Please rank from very concerned to very comfortable, or n/a.**

very

concerned

slightly

concerned

neither

concerned or comfortable

somewhat

comfortable

very

comfortable

n/a

Unexplained episode of DKA in	the last 12 months

HbA1c at or close to target in	current pump user

HbA1c consistently >10%	Infrequent glucose monitoring	Erratic schedule/meal pattern

Evidence of depression and/or	diabetes related distress

Obsessive compulsive	tNeontduesnincigesbolus calculator - no

set insulin: carb, or insulin sensitivity factor

28. Are there any other factors that you would	Yes

consider to be important/essential prerequisites for	No

AID use?	Unsure

If yes, what would these be?

If no, why is this?

If unsure, why is this?

29. Do you consider yourself to be actively involved	Yes

in the care of patients using DIY AID systems?	No Unsure

29. a) When you review your patient who is using DIY	never AID do you review their blood glucose data through the	rarely

specific app they are using (xDrip, Glimp, Spike etc)?	sometimes

frequently always n/a

29. b) How comfortable do you feel in doing this?	not at all

somewhat uncomfortable

neither comfortable or uncomfortable comfortable

very comfortable n/a

29. c) If your patient is using DIY AID, do you	never

discuss with them any suggestions for alterations to	rarely

their settings?	sometimes

frequently always n/a

29. d) How comfortable do you feel in doing this?	not at all

somewhat uncomfortable

neither comfortable or uncomfortable comfortable

very comfortable n/a

29. e) If your patient is using DIY AID, do you	never

discuss with them any specific support platforms	rarely

(social media, online communities) they are using?	sometimes frequently always

n/a

29. f) How comfortable do you feel in doing this?	not at all

somewhat uncomfortable

neither comfortable or uncomfortable comfortable

very comfortable n/a

**30. How much would the following make you more confident to recommend and support a patient using a COMMERCIAL AID system?**

**Please rank from not at all to yes definitely, or n/a.**

not at all	probably not	unsure	yes possibly	yes definitely	n/a

Clinical Practice Guidelines from	Diabetes Canada

An educational program for	uAsneersducational program for HCPs

Support for the USER from a	certified HCP

Support for YOU from a certified	HCP

Medico-legal guidance (eg	consent forms, checklist,

waivers)

Are there any other suggestions that would make you

more confident to recommend and support a patient	using a Commercial AID system?

**31. How much would the following make you more confident to recommend and support a patient using DIY AID system?**

**Please rank from not at all to yes definitely, or n/a.**

not at all	probably not	unsure	yes possibly	yes definitely	n/a

Clinical Practice Guidelines from	Diabetes Canada

An educational program for	uAsneersducational program for HCPs

Support for the USER from a	certified HCP

Support for YOU from a certified	HCP

Medico-legal guidance (eg	consent forms, checklist,

waivers)

Are there any other suggestions that would make you

more confident to recommend and support a patient	using a DIY AID system?
